# A Vital Sign-based Prediction Algorithm for Differentiating COVID-19 Versus Seasonal Influenza in Hospitalized Patients

**DOI:** 10.1101/2021.01.13.21249540

**Authors:** Naveena Yanamala, Nanda H. Krishna, Quincy A. Hathaway, Aditya Radhakrishnan, Srinidhi Sunkara, Heenaben Patel, Peter Farjo, Brijesh Patel, Partho P Sengupta

## Abstract

Patients with influenza and SARS-CoV2/Coronavirus disease 2019 (COVID-19) infections have different clinical course and outcomes. We developed and validated a supervised machine learning pipeline to distinguish the two viral infections using the available vital signs and demographic dataset from the first hospital/emergency room encounters of 3,883 patients who had confirmed diagnoses of influenza A/B, COVID-19 or negative laboratory test results. The models were able to achieve an area under the receiver operating characteristic curve (ROC AUC) of at least 97% using our multiclass classifier. The predictive models were externally validated on 15,697 encounters in 3,125 patients available on TrinetX database that contains patient-level data from different healthcare organizations. The influenza vs. COVID-19-positive model had an AUC of 98%, and 92% on the internal and external test sets, respectively. Our study illustrates the potentials of machine-learning models for accurately distinguishing the two viral infections. The code is made available at https://github.com/ynaveena/COVID-19-vs-Influenza and may be have utility as a frontline diagnostic tool to aid healthcare workers in triaging patients once the two viral infections start cocirculating in the communities.

## Main

Infection with severe acute respiratory syndrome coronavirus 2 (SARS-CoV 2) causing coronavirus disease 2019 (COVID-19) has led to an unprecedented global crisis due to its vigorous transmission, spectrum of respiratory manifestations, and vascular affects ^1-3^. The etiology of the disease is further complicated by a diverse set of clinical presentations, ranging from asymptomatic to progressive viral pneumonia and mortality ^4^. Due to its similar symptomatology, COVID-19 has drawn comparisons to the seasonal influenza epidemic.^5^ Both infections commonly present with overlapping symptoms, leading to a clinical dilemma for clinicians as SARS-CoV 2 carries a case-fatality rate up to 30 times that of influenza and infects healthcare workers at a significantly higher rate.^3,6,7^ Moreover, the concurrence of epidemics appears imminent as the considerable COVID-19 incidence continues and even a moderate influenza season would result in over 35 million cases and 30,000 deaths.^5,6^ To help curb this dilemma, front-line providers need the ability to rapidly and accurately triage these patients.

One approach to quickly classifying patients as COVID-19 positive or negative could be through machine learning algorithms. While the use of machine learning has been applied to contact tracing and forecasting during the COVID-19 epidemic ^8^, it has only limitedly been explored as a means for accurately predicting COVID-19 infection on clinical presentation. With just a few important parameters clinicians can diagnose the patients well before a laboratory diagnosis. Preliminary work has shown the utility of machine and deep learning algorithms in predicting COVID-19 for patient features ^9-11^ and on CT examination ^12,13^, but there remains a paucity in research showing the capacity of machine learning algorithms in differentiating between COVID-19 and influenza patients.

Vital signs are critical piece of information used in the initial triage of patients with COVID-19 and/or influenza by care coordinators and health-care responders in community urgent care centers or emergency rooms. It is becoming clearer that patient vital signs may present uniquely in SARS-CoV 2 infection ^9^, likely as a result of alterations in gas exchange and microvascular changes ^14^. In the present investigation, we therefore explored the use of machine learning models to differentiate between SARS-CoV 2 and influenza infection using basic office-based clinical variables. The use of simple ML-based classification may have utility for the rapid identification, triage, and treatment of COVID-19 and influenza positive patients by front-line healthcare workers, which is especially relevant as the influenza season approaches.

## Results

### WVU Study Cohort

#### Baseline Characteristics

The patient cohort included 3883 patients (mean age 52 ± 24 years, 48% males, and 89% White/Caucasian) of whom 747 (19%) tested positive for SARSCoV-2 (COVID-19 positive cohort), 1913 (49%) tested negative for SARSCoV-2 (COVID-19 negative cohort), and 1223 (31%) had influenza (Table 1, Figure S1). The majority of the COVID-19 positive and negative patients were older; whereas the influenza cohort was younger (P<0.001). There was higher prevalence of Black/African Americans in the influenza cohort in comparison with other cohorts. COVID-19 positive patients were more obese (p<0.001). They also had higher mean body temperature compared to COVID-negative patients and exhibited an overall higher systolic and diastolic blood pressures than the other two cohorts (p<0.001 for all variables). While the Influenza cohort had higher mean body temperature, heart rate and oxygen saturations than COVID-19-positive and -negative patients (p<0.001). Patients in the COVID-19-positive and influenza groups had a higher respiratory rate than the COVID-19-negative group (Table 1).

**TABLE 1.** Demographics of WVU Patients (included COVID-19-positive, COVID-19-negative and Influenza cohort)

|  | Overall | COVID-19-positive | Influenza | COVID-19-negative | p value |
| --- | --- | --- | --- | --- | --- |
| **N** | 3883 | 747 | 1223 | 1913 |  |
| **Overall Age, in years** | 52 ± 24 | 55 ± 22<sup>*</sup> | 41 ± 24<sup>†</sup> | 57 ± 23 | < 0.001<sup>‡</sup> |
| **Age by groups** |  |  |  |  | < 0.001<sup>§</sup> |
| Kids & Teens (0-20) | 520 | 63 (8.4) | 319 (26.1) | 138 (7.2) |  |
| Adults (21-44) | 940 | 171 (22.9) | 349 (28.5) | 420 (22.0) |  |
| Older Adults (45-64) | 1005 | 216 (28.9) | 282 (23.1) | 507 (26.5) |  |
| Seniors (65 or older) | 1418 | 297 (39.8) | 273 (22.3) | 848 (44.3) |  |
| **Male, n(%)** | 1876 | 375 (50.2) | 585 (47.8) | 916 (47.9) | 0.51 |
| **Race** |  |  |  |  | < 0.001<sup>§</sup> |
| White/Caucasian, n(%) | 3455 | 641 (85.8) | 1037 (84.8) | 1777 (92.9) |  |
| Black/African-American, n(%) | 203 | 48 (6.43) | 95 (7.8) | 60 (3.1) |  |
| Other, n(%) | 225 | 58 (7.8) | 91 (7.4) | 76 (4.0) |  |
| **Ethnicity** |  |  |  |  | < 0.001<sup>§</sup> |
| Hispanic/Latino, n(%) | 94 | 34 (4.6) | 43 (3.5) | 17 (0.9) |  |
| Other, n(%) | 3789 | 713 (95.5) | 1180 (96.5) | 1896 (99.1) |  |
| **Body Mass Index, kg/m<sup>2</sup>** | 30.0 ± 9.4 | 31.6 ± 8.8<sup>*†</sup> | 29.7 ± 9.5 | 29.6 ± 9.5 | < 0.001<sup>‡</sup> |
| **Vitals, (mean±SD)** |  |  |  |  |  |
| Temperature in °F, | 98.3 ± 1.1 | 98.3 ± 1.1<sup>*†</sup> | 99.0 ± 1.3<sup>†</sup> | 98.0 ± 0.8 | < 0.001<sup>‡</sup> |
| Systolic blood pressure, mmHg | 126 ± 20 | 127 ± 19<sup>*</sup> | 124 ± 19<sup>†</sup> | 126 ± 20 | 0.004<sup>‡</sup> |
| Diastolic blood pressure, mmHg | 73 ± 13 | 75 ± 13<sup>*†</sup> | 72 ± 12 | 73 ± 13 | < 0.001<sup>‡</sup> |
| Heart rate, beats per minutes | 87 ± 18 | 83 ± 16<sup>*</sup> | 94 ± 20<sup>†</sup> | 83 ± 17 | < 0.001<sup>‡</sup> |
| Oxygen Saturation (SpO2), % | 96 ± 4 | 96 ± 4<sup>*†</sup> | 97 ± 3<sup>†</sup> | 96 ± 4 | < 0.001<sup>‡</sup> |
| Respiratory rate | 19 ± 4 | 19 ± 4<sup>*</sup> | 19 ± 3<sup>†</sup> | 18 ± 5 | < 0.001<sup>‡</sup> |

Values reported are counts (%) or mean ± standard deviation.
\* p < 0.05 compared with the influenza group.
† p < 0.05 compared with the COVID-19-negative group.
‡ p < 0.05 using Kruskal Wallis test with Dunn-Bonferroni correction.
§ p < 0.05 using Chi-square test.

#### Outcomes

The overall mortality for the cohort was 6.7%. The crude case fatality rate was 6.8% in the COVID-19-positive and 4.2% in influenza groups, with a 9.5% case fatality rate in the COVID-19 negative group (p<0.001). The COVID-19-positive patients had more than a 3-fold higher rate for ICU admissions than patients with influenza (19.0% vs. 5.7%; p<0.001), but the rate was lower than the COVID-19 negative group (23.2%) (p<0.001). The average age of patients who died during hospitalizations was significantly higher in COVID-19 positive patients (75 ± 14 years) compared to the influenza (69 ± 13years) and COVID-19 negative groups (72 ± 14 years) (p=0.02), as presented in the Table 2.

**TABLE 2.**
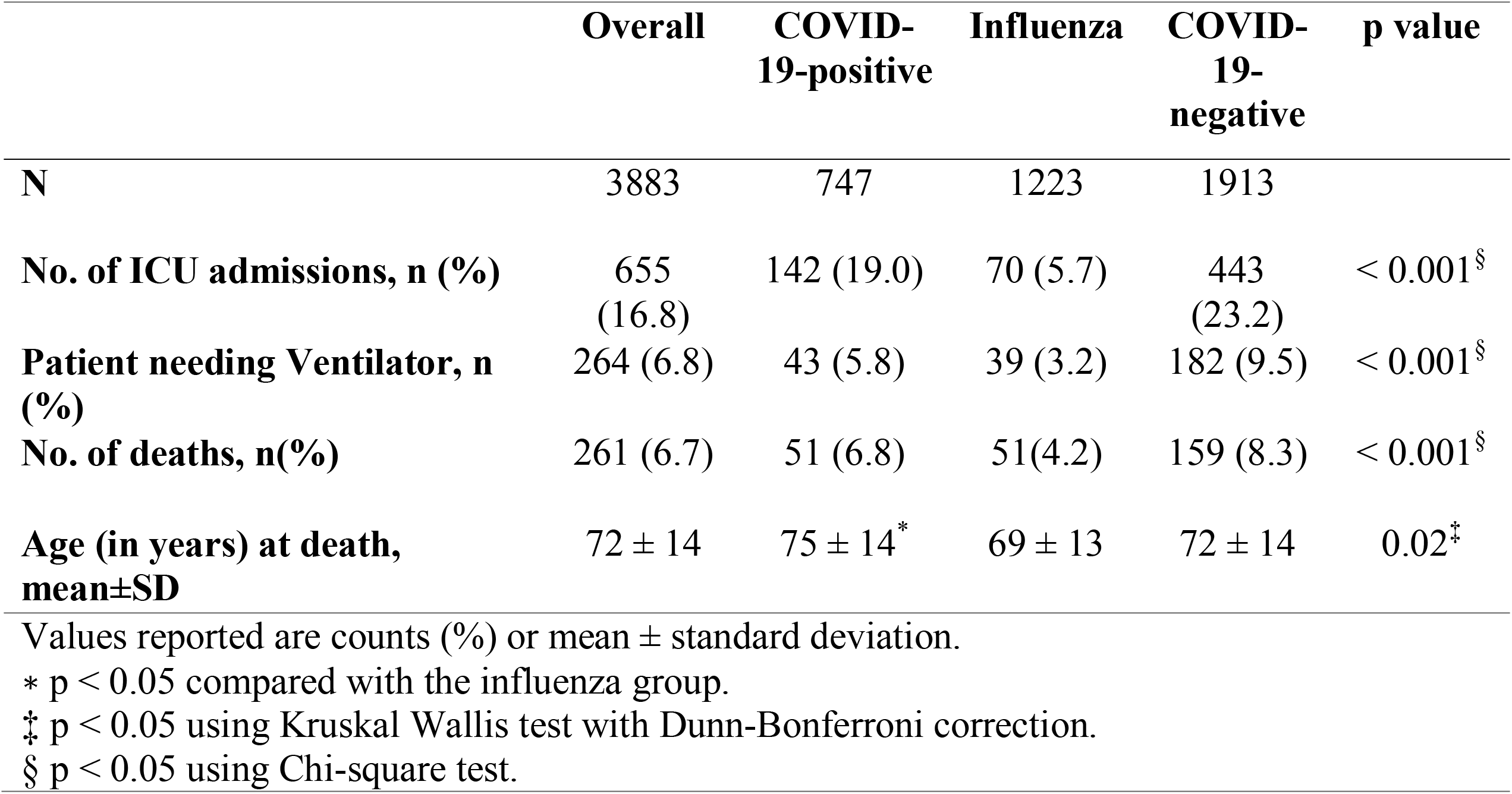
Outcomes of WVU Patients presented at ED or admitted to WVU hospitals and tested positive for COVID-19, positive for Influenza A/B or negative for COVID-19.

### TriNetX Cohort

The external cohort included a total of 15,697 patient encounters from 3.125 patients with body temperature information available (Supplementary Table 2). This subgroup of the external cohort included 6,613 encounters from 1,057 COVID-positive patients and 9,087 encounters from 2,068 influenza patients. The COVID-19-positive group was predominantly male (54%), while the influenza group involved more female (53%) patients. The COVID-19-positive group included more Black/African-American (47%) patients, while the influenza cohort consisted of more White/Caucasian (52%) patients. The vital signs for mean body temperature, heart rate, respiratory rate, and diastolic blood pressure showed a statistical difference between patients with COVID-19 and those with influenza (p <0.0001).

### COVID-19 Versus Influenza Infection Prediction at the ED or Hospitalization

In this study we explored the value of demographics, vitals and symptomatic features, which are readily available to providers, in an effort to develop supervised machine learning classifiers that can predict patients who are either COVID-19 positive or negative, while further distinguishing influenza from COVID-19 infection. The WVU hospitalized patient cohort was randomly divided into a training (80%) and testing set (20%) to develop four different contexts specific XGBoost predictive models. We assessed receiver operator characteristic (ROC) area under the curve (AUC) plots, precision, recall and other threshold evaluation metrics to select the best performing model in each case (Tables 3, 4 & S3).

**TABLE 3.**
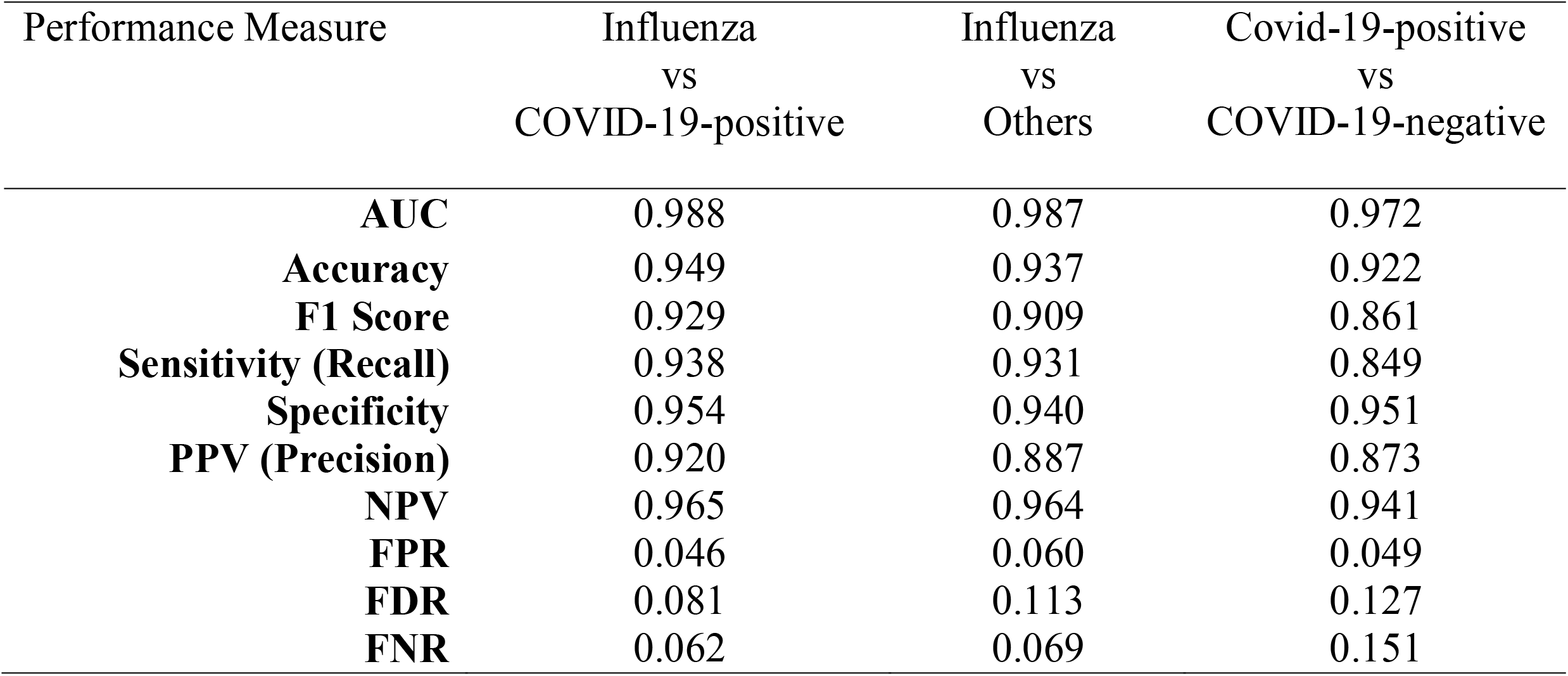
Performance metrics of different XGBoost models for predicting the given record as COVID-19-positive or influenza when tested on Internal validation or test set.

| Performance Measure | Influenza<br>vs<br>COVID-19-positive | Influenza<br>vs<br>Others | Covid-19-positive<br>vs<br>COVID-19-negative |
| --- | --- | --- | --- |
| **AUC** | 0.988 | 0.987 | 0.972 |
| **Accuracy** | 0.949 | 0.937 | 0.922 |
| **F1 Score** | 0.929 | 0.909 | 0.861 |
| **Sensitivity (Recall)** | 0.938 | 0.931 | 0.849 |
| **Specificity** | 0.954 | 0.940 | 0.951 |
| **PPV (Precision)** | 0.920 | 0.887 | 0.873 |
| **NPV** | 0.965 | 0.964 | 0.941 |
| **FPR** | 0.046 | 0.060 | 0.049 |
| **FDR** | 0.081 | 0.113 | 0.127 |
| **FNR** | 0.062 | 0.069 | 0.151 |

**TABLE 4.**
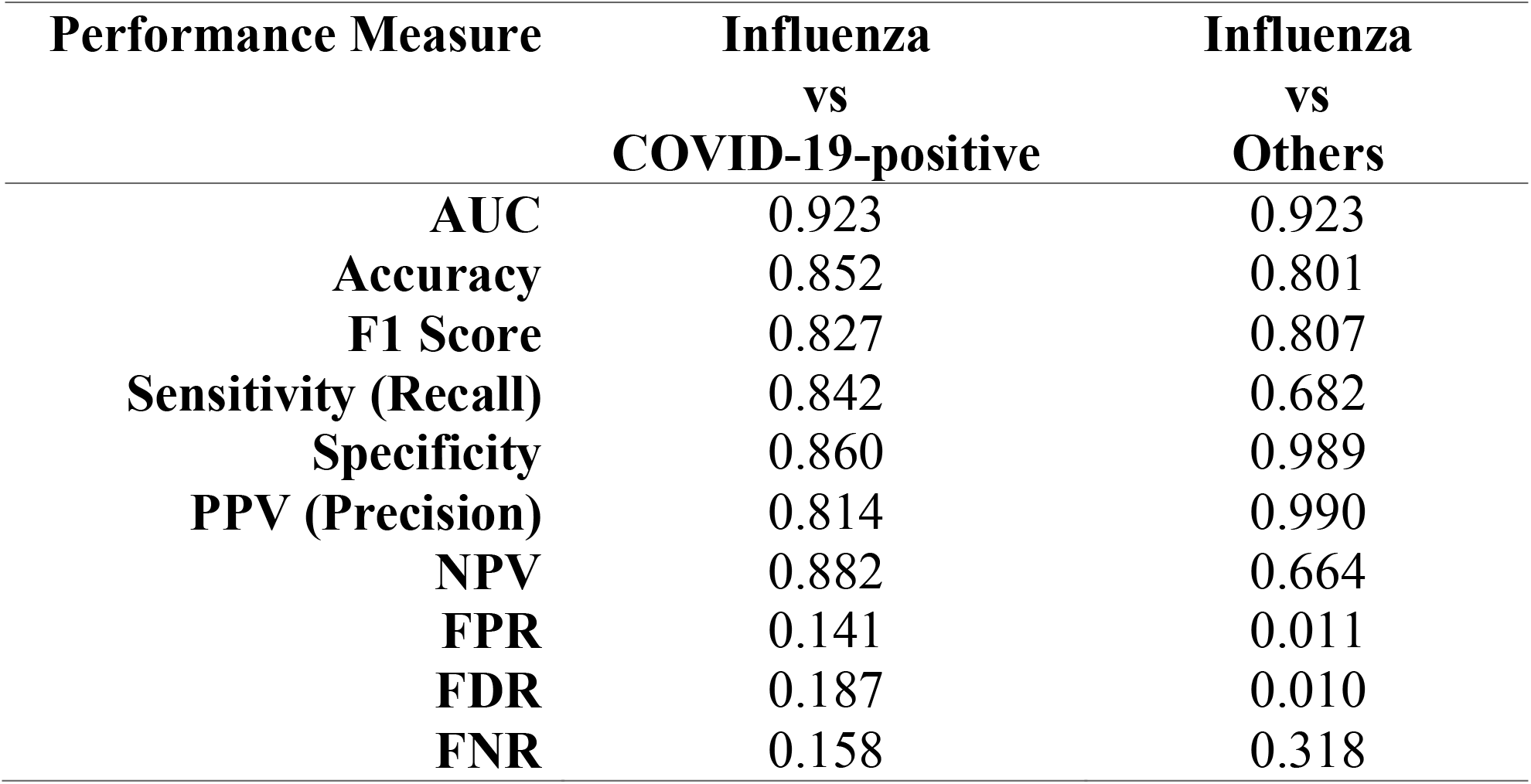
Performance of Influenza versus COVID-19-positive and Influenza versus Others (COVID-19-positive/-negative) classification models on the external validation cohort.

| **Performance Measure** | **Influenza**<br>**vs**<br>**COVID-19-positive** | **Influenza**<br>**vs**<br>**Others** |
| --- | --- | --- |
| **AUC** | 0.923 | 0.923 |
| **Accuracy** | 0.852 | 0.801 |
| **F1 Score** | 0.827 | 0.807 |
| **Sensitivity (Recall)** | 0.842 | 0.682 |
| **Specificity** | 0.860 | 0.989 |
| **PPV (Precision)** | 0.814 | 0.990 |
| **NPV** | 0.882 | 0.664 |
| **FPR** | 0.141 | 0.011 |
| **FDR** | 0.187 | 0.010 |
| **FNR** | 0.158 | 0.318 |

Panel A and B in Figure 1 shows the ROC curves for the prediction of influenza from COVID-19 positive encounters and from those who are either COVID-19 positive or negative obtained using the holdout test set. We present four unique models that can be used as a framework to aid in the delineation between influenza and COVID-19 in the clinical setting. The first model provided stratification of patients as either influenza or COVID-19 positive, highlighted by a ROC AUC 98.8%, accuracy of 95%, sensitivity 94%, and specificity 95% at identifying COVID-19-positive patients (Table 3). The second model distinguished influenza patients from all other patients, irrespective of a patient’s COVID-19 test, revealing an ROC AUC of 98.7%, accuracy of 94%, sensitivity of 93%, and specificity of 94% (Figure 1, Panel B). The third model distinguishes between COVID-19 positive and negative patients, with a ROC AUC of 97.2%, accuracy of 92%, sensitivity of 85%, and specificity of 95% (Figure 2, Panel A and Table 3). The Precision-Recall AUC was 94% for predicting COVID-19 positive versus negative patients (Figure S3).

**Figure 1.**
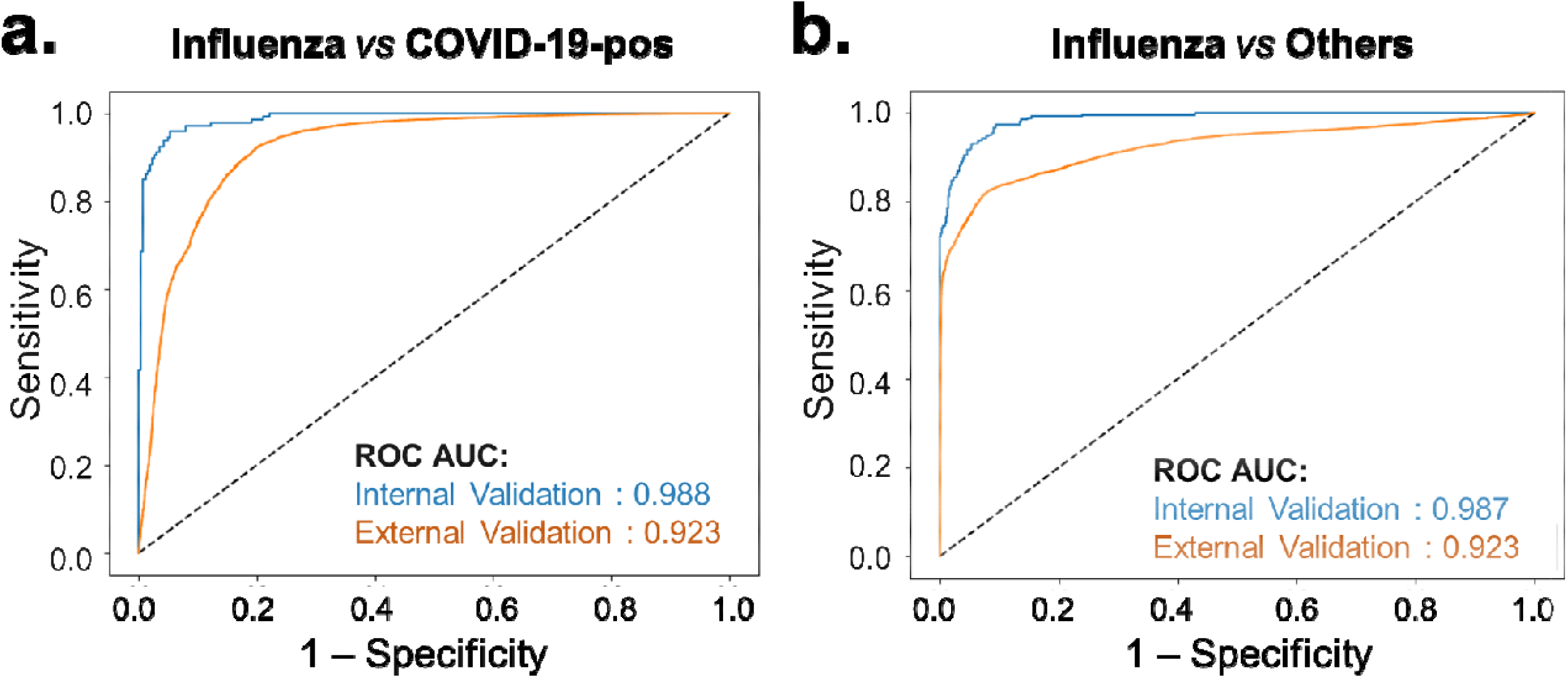
Receiver Operating Characteristic Curves showing the predictive performance of the (a) influenza versus COVID-19-positive model and (b) Influenza versus Other prediction models on the internal test and external validation datasets.

The fourth model employed a multi-class XGBoost framework trained to distinguish between all three different types of patients had an ROC AUC of 98% and achieved 91% precision at identifying patients that were positive for influenza with 91% recall, and 95% precision at identifying those patients that tested negative for COVID-19 with a 88% recall. While the highest specificity with precision was achieved when identifying COVID-19 positive patients, the recall of 75% was significantly lower compared to other classes (91% for influenza and 88% for COVID-19 negative patients). Average precision of the overall model was 91%, accuracy was 90% and macro-average of ROC AUC was 97.6% (Figure 2 and Table S3).

**Figure 2.**
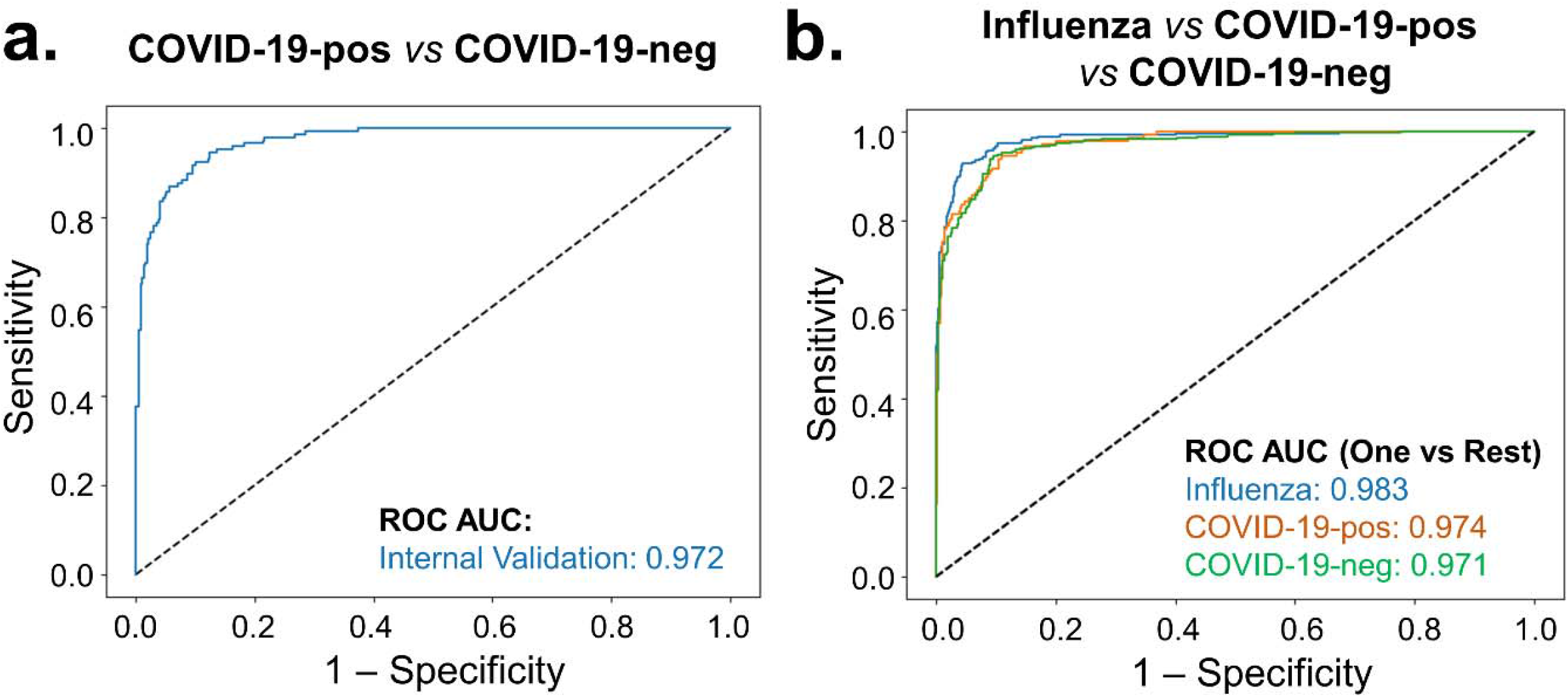
Receiver Operating Characteristic Curves showing the predictive performance of the (a) COVID-19-positive versus COVID-19-negative, and (b) Influenza versus Other prediction models on the internal validation test.

### Importance of Various Features

We use the SHapley Additive exPlanations (SHAP) method ^15,16^, specifically, using the Tree Explainer method, to describe our XGBoost models.

#### SHAP Importance

SHAP is a model-agnostic interpretability method that aids in analyzing feature importance based on their impact on the model’s output. The additive importance of each feature for the model is calculated over all possible orderings of features. Positive SHAP values indicate a positive impact on the model’s output, while negative values indicate negative impact on the model’s output. The most important features that identify patients with positive influenza infection from others that are presented to ED included features such as age, body temperature, body surface area (BSA) and heart rate (Figure 3A and B). Encounter-related features such as the month of encounter along with reason for visit (Table S4) and encounter type also contributed to the most-informative variables for predicting influenza compared to COVID-19 positive patients or influenza vs all other patients (Figure 3). On the contrary, in the case of the COVID-19-positive vs COVID-19 negative model, two vital signs i.e., body temperature and SPO_2_ were amongst the highest-ranking features. Age, BMI, diastolic blood pressure and encounter-related variables such as reason for visit and month of encounter were additionally amongst the variables most informative to model predictions. A similar trend of feature importance was also reflected in the SHAP summary plot of the three-way multi-class classifier (Figure S2). Vital signs played a more significant role in distinguishing between influenza and COVID-19 positive encounters through parameters such as body temperature, heart rate and blood pressure.

**Figure 3.**
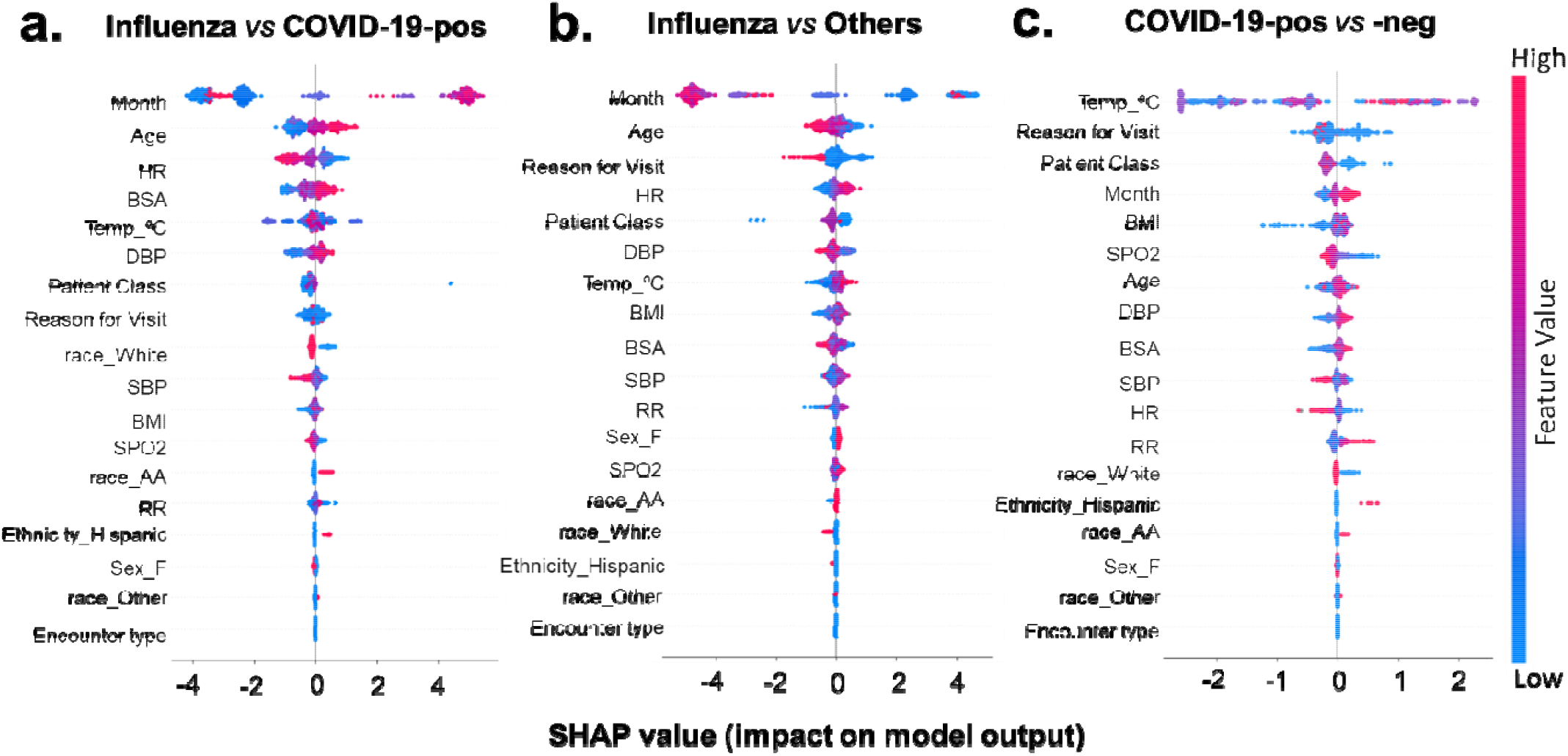
SHapley Additive exPlanations (SHAP) beeswarm summary plot of shap values distribution of each feature of the test dataset. The plot depicts the relative importance, impact and contribution of different features on the output of (a) Influenza vs COVID-19-positive, (b) Influenza vs Other and (c) COVID-19-positive vs COVID-19-negative predictive models. The summary plot combines feature importance with feature effects. The features on the y-axis are ordered according to their importance. Each point on the summary plot is a Shapley value for a feature and an instance (i.e., a single patient encounter in this case) in the dataset. The position of each point on the x-axis shows the impact that feature has on the classification model’ prediction for a given instance. The color represents the high (red) to low (blue) values of the feature (i.e., Age, BMI etc.).

From the SHAP summary plots (Figure 3 & S2), it is evident that the models captured some important features and patterns that aid in correctly predicting Influenza and COVID-19-positive encounters. These plots highlighted that patient encounters predicted to be COVID-19 positive are, on average, more likely to have lower heart rate, higher respiratory rate and lower oxygen saturation. Lower blood oxygen saturation is known to be prevalent among COVID-19 patients and was identified by our machine learning models as an important feature for validating the clinical appropriateness of our model design.

#### Model Interpretability

The SHAP force plots shown in Figures 4 & 5 aim to offer explanations for individual predictions made by our models. The two plots generated for the COVID-19-positive vs influenza classifier are shown in Figure 4. Panel A shows an encounter correctly classified as influenza, with the month (during the time course of the traditional influenza season), BSA and age (younger patient) values affecting the model output most. The effect of these features was enough to prevent misclassification due to lower heart rate and SBP values. Panel B explains an encounter correctly classified as COVID-19. While the encounter occurred in month 3, which overlaps with the influenza season, the combined effect of higher age, low heart rate, BSA and higher temperature pushes the prediction towards COVID-19, counteracting the effect of month and encounter type.

**Figure 4.**
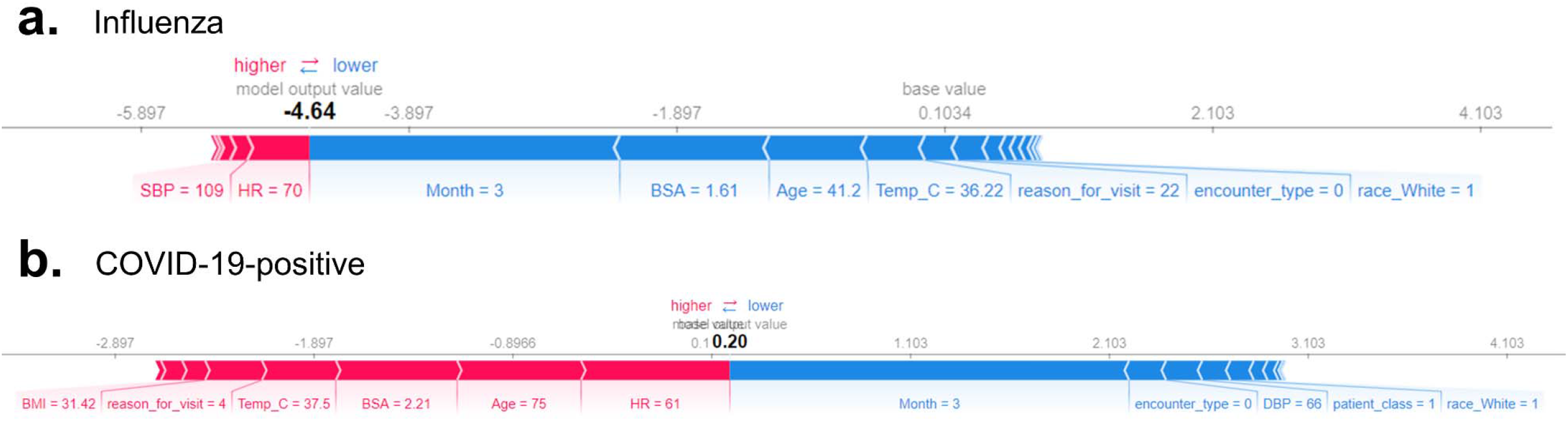
SHapley Additive exPlanations (SHAP) force plots for sample observations from (a) Influenza A/B and (b) COVID-19-positive predictions. The underlying model is XGBoost. Features that are contributing to a higher and lower SHAP values are shown in red and blue, respectively along with the size of each feature’s contribution to the classification model’s output. The influenza patient in this example shows a log-odds output of -4.64 in the rating scale, which is equal to a probability of 0.0096. The baseline – the mean of the model output (log-odds) over the training dataset – is 0.1034 (translating to a probability of 0.5258). The COVID-19 positive patient has a rating score of 0.20 (probability = 0.5498); Age of 75, HR of 61, BSA of 2.21 and a Temp of 37.5°C increases the prediction risk, while Month of march decreases the predicted risk of being COVID-19 positive.

**Figure 5.**
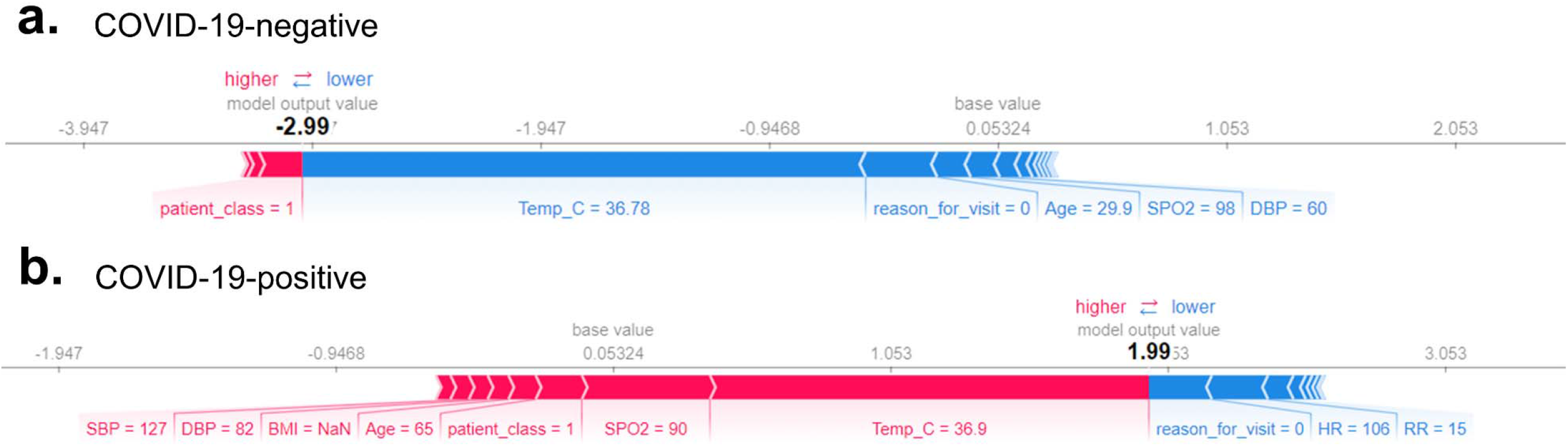
SHapley Additive exPlanations (SHAP)force or explanation plots of two patient encounters with (a) COVID-19-negative and (b) COVID-19-positive predictions. Features that are contributing to a higher and lower SHAP values are shown in red and blue, respectively along with the size of each feature’s contribution to the model’s output. The baseline -- the mean of the model output (log-odds) over the training dataset -- is 0.053 (translating to a probability value of 0.5132). The first patient – who is COVID-19 negative – has a low predicted risk score of -2.99 (output probability = 0.0479). The second patient – who is COVID-19 positive – has a high predicted risk of 1.99 (output probability = 0.8797). These SHAP output values represent a ‘raw’ log-odds value which is transformed into a probability space, to provide the final output of 0 and 1 (< 0.5 and > 0.5). Risk increasing effects such as Temp, SpO_2_, Age and BP were offset by decreasing effects of HR and RR in pushing the model’s predictions towards or away from the positive class, respectively.

Similarly, a second set of plots were generated for the COVID-19 positive vs negative classifier (Figure 5). In these plots, Panel A shows an encounter correctly classified as COVID-19 negative while Panel B shows an encounter classified correctly as COVID-19 positive. We see in the former that the temperature, reason for visit, lower age and higher value of SPO_2_ push the prediction towards COVID-19 negative. In the latter, we see that the effect of temperature, low SPO_2_ value and higher age together counter the effect of the higher heart rate and normal respiratory rate, to correctly classify the example as COVID-19 positive. With these interpretability methods we are able to clearly determine the reasons for the model’s output and ensure they can be scrutinized. The insights obtained further corroborate with patterns often observed in COVID-19 patients.

### Impact of Vitals on Model Performance

#### Stepwise removal of vitals based on feature importance

Each feature’s importance to the construction of the machine learning model was assessed through individually removing each vital sign parameter in the internal validation set. Stepwise removal of vitals in the order of their importance of contribution to the multi-class classifier (Figure S2) led to decrease in the performance of the model (Figure S4A). Removal of body temperature significantly decreased models’ performance to an accuracy of 74% and F1 score of 68% compared to the initial performance 90% and 89%, respectively. However, subsequent removal of other vital signs, including heart rate did not affect model performance drastically.

#### Inclusion of only one vital sign at a time

We also assessed how well the model performs given only one vital is present at a time in the Internal test set. More specifically, when considering the importance of a given vital sign, all other features related to vitals are removed from the internal test set. However, the demographics and other information was not ablated. Figure S4B shows the models performance in terms of F1 score or statistic by only including one vital at a time. The higher the performance, the more useful the vital sign is in helping the model to discriminate between COVID-19 and influenza encounters. With the comparison of both multi-class classifier and COVID-19 positive vs negative classifier performances, it is evident that body temperature has the greatest impact among all vital signs considered. In addition to body temperature, heart rate was also seen to contribute to the performance of COVID-19 positive vs negative model.

Taken together, these results suggest that of all the vital signs, body temperature, followed by heart rate and SPO_2_ could impact the predictive models’ performance in discriminating between influenza, COVID-19 positive and negative patient encounters.

### External Validation

To further assess the generalizability of the predictive models and confirm the stability of the model features at identifying patients positive for COVID-19 or influenza, we validated our models using the TriNetX research network dataset external to WVU Medicine. Patients with any missing data related to body temperature were excluded from the analysis. The dataset included 6,613 encounters of COVID-19 patients (n=1,057) and 9,084 encounters related to influenza patients (n=2,068). The influenza versus COVID-19 model demonstrated ROC AUC of 92.3% with an accuracy of 85%, and 81% precision at identifying patients that were positive for COVID-19 with 84% recall (Figure 1A & Table 4). Also, the model to detect patients with influenza, irrespective of their COVID-19 status, showed similar performance with an AUC ROC of 92.3% and an accuracy of 80% %, and 99% precision at identifying patients that were positive for influenza with 68% recall (Figure 1B). This suggests that the developed models are able to effectively identify patients across multiple TriNetX HCOs with influenza or COVID-19 infections amongst other COVID-19-negative patients presenting at ED and/or admitted to the hospital.

Further, enforcing no missing values in the case of both heart rate and body temperature, the two top ranked vital signs (Figure S4), did not result in boosting the performance. On the contrary, restricting the data to have all vitals present, boosted the performance of ROC AUC to and accuracy to 88% for the Influenza vs COVID-19 model and ROC AUC to and accuracy to in the case of Influenza vs Other model (Table S5). These results suggest that while enforcing no missing values in vitals could support a better overall model performance (i.e., AUC ROC 94.3% vs 92.3%), missingness in most of the vitals does not seem to limit its applicability and generalizability.

## Discussion

This investigation provides multiple machine-learning models to differentiate between COVID-19 positive, -negative and influenza. Further, this is the first machine learning model to leverage a patient population that includes both the initial (February-April) and secondary (May-September) surge of SARS-CoV 2 infections in the United States ^17^. Our approach can guide future applications, highlighting the importance of developing dynamic models that control for confounding comorbidities, such as influenza, and the ever-evolving infectivity of SARS-CoV 2. Importantly, as influenza season approaches it will be of high priority to establish a reliable process for identifying patients at risk of COVID-19, influenza, or other viral infections to increase the prognostic value of a directed therapy.

While the initial presentation of COVID-19 and influenza appear similar, the number, and combination, of signs and symptoms can help provide better stratification. From a symptom standpoint, COVID-19 causes more fatigue, diarrhea, anosmia, acute kidney injury, and pulmonary embolisms, while sputum production and nasal congestion are more specific to influenza ^18^. Furthermore, COVID-19 tends to cause worse decompensation. For instance, out of 200 cases of either influenza or COVID-19 in a study from France, only COVID-19 resulted in severe respiratory failure requiring intubation^18^. Our evaluation revealed that the month the patient was seen, age, and heart rate were the most important features for predicting a diagnosis of COVID-19 over that of influenza infection. Additionally, body temperature and SPO_2_ were two of the most important features that indicated if a patient would be positive or negative for COVID-19, which is likely a result of the underlying pathophysiology of the virus. SARS-CoV 2 infection appears to cause more significant endothelial dysfunction and systemic inflammation likely leading to the worse case-fatality rate.^6,19^ These differences can be seen histologically as well. Autopsy samples of COVID-19 lung tissue showed severe endothelial injury and widespread thrombosis with microangiopathy ^20^. These samples had nine times as much alveolar capillary microthrombi compared to that of influenza lung tissue and even showed evidence of new vessel growth through a mechanism of intussusceptive angiogenesis ^1,20^.

Machine learning algorithms can provide distinct advantages in the classification of positive COVID-19 cases as the virus, and the demographic it infects, continues to evolve. The West Virginia University (WVU) cohort consists of a fairly homogenous population, with 89% of the population identifying as White/Caucasian and only 5% as Black/African American. While our machine learning models were built around the WVU cohort, they demonstrate robust performance even on more racially/ethnically diverse populations, such as in the TriNetX dataset, when predicting patients who have influenza or COVID-19 (ROC AUC = 94.3%) and influenza only (ROC AUC = 96.2%). The generalizability of our model may suggest that features that define SARS-CoV 2 infection (e.g., age, heart rate, body temperature, SPO_2_) could represent similarities in clinical presentation, regardless of a patient’s race/ethnicity. Further understanding the socioeconomic ^21,22^ and physiological ^23,24^ differences in minority populations will be important in determining why COVID-19 disproportionately affects these populations and how machine learning models can more accurately model heterogeneous populations.

Patient information provided in the WVU and TriNetX datasets consists of both the initial rise in cases in the United States (February-April) as well as a secondary surge (May-September). The second surge of infections shifted demographically from a predominately older population to one with the highest prevalence between ages 20-29, with an increased rate of mild to asymptomatic presentations ^25^. Additionally, SARS-CoV 2 is an RNA virus with the capacity to mutate. Sequencing studies have already identified a variety of genetic variants, with most mutations currently having no clear association with a positive or negative selective virulence ^26^. Though, some mutations, such as D614G, are suggested to increase infectivity of SARS-CoV 2 and outcompete other strains in the environment ^27^. Regardless, in our current approach, applying machine learning over the continuum of the COVID-19 epidemic provides distinct advantages in producing a generalizable model for continued use, which may not be captured in models generated on earlier datasets.

The current study provides robust ROC AUC prediction of patients with COVID-19 (98.3%), without COVID-19 (97.4%), and those with influenza (97.1%) in our multi-class model. While only demographic information and vital signs were applied to our models, overall prediction could likely improve with the addition of other features including biochemical, metabolic, and molecular markers, as well as imaging modalities such as X-ray and CT. The addition of these features could likely improve diagnostic accuracy and should be tested in future studies delineating between COVID-19 and influenza. Additionally, the current investigation involves influenza and COVID-19 data from non-overlapping periods (i.e., influenza from winter 2019 and COVID-19 from spring/summer/fall 2020). Although some of the machine learning models rely on time of presentation for prediction, we anticipate that the four-model framework provided in the study will sufficiently stratify patients even when influenza season and the COVID-19 epidemic overlap. This is reinforced by the prediction of COVID-19 positive and negative cases (ROC AUC = 97.2%) in our model, which did not rely on time of presentation as a variable.

Our study was limited by the classification of patients in the internal (WVU) and external (TriNetX) datasets. While the WVU cohort includes patients, who have been confirmed to have a negative SARS-CoV 2 test on presentation, the TriNetX dataset does not have this information. Both datasets are also unable to capture patients with other respiratory viruses and viral coinfections; information that could better explain variations in COVID-19 clinical presentation. Another limitation is the need to continually adjust these models to fit new trends in SARS-CoV 2 spread and infectivity. Our models benefit from the inclusion of data spanning from February to October, which can better simulate the current COVID-19 epidemic and influenza season but will still require future iterations and validations to accurately predict SARS-CoV 2 infections. Further, an understanding of COVID-19 and viral co-infections is needed to appropriately model the risks of patients presenting with both illnesses ^28-30^.

## Conclusions

Here we highlight how machine learning can effectively classify influenza and COVID-19 positive cases through vital signs on clinical presentation. This work is the first step in building a low-cost, robust classification system for the appropriate triage of patients displaying symptoms of a viral respiratory infection. With these algorithms, the identification of proper treatment modalities for both COVID-19 and influenza can be made more rapidly, increasing the effectiveness of patient care.

## Outlook

We certainly hope that our current work can aid healthcare workers and clinicians to rapidly identify, triage, and guide treatment decisions when the two viral infections start cocirculating in the communities. In addition, Further studies validating the model as we see more coinfection in an upcoming season can significantly improve the quality of care and proper allocation of resources during this global pandemic.

## Materials and Methods

### WVU Internal Cohort Dataset

De-identified clinical and demographics data for all patients presenting to the emergency department and/or admitted at West Virginia University hospitals between January 1^st^, 2019, and November 4^th^, 2020, were extracted and provided to us by WVU Clinical and Translational Science Institute (CTSI). The COVID-19 positive dataset includes all patients who had a lab-based diagnostic test positive for SARS-CoV-2 at WVU Medicine between March 1^st^, 2020, and November 4^th^, 2020 and had an associated hospital encounter with all vital signs information available. The influenza positive dataset includes a random sample of 1500 patients who had a lab-based diagnostic test that was positive for either Influenza Type A or Influenza Type B at WVU Medicine between January 1st, 2019, and November 3rd, 2020, and had a hospital encounter associated with that positive test (Figure S1). The COVID-19 negative dataset includes a random sample of 2000 patients who had negative test results for SARS-CoV-2 and who never had a positive diagnostic test for SARS-CoV-2 at WVU Medicine between March 1^st^, 2020, & November 4^th^, 2020 provided that an associated hospital encounter occurred with that negative test. As our study aims to create a model that can accurately discriminate COVID-19 from influenza patients solely based on the patient’s demographic information and vital signs data, patients with any of the vital signs missing during the associated encounters were therefore excluded from the analysis. This resulted in a final WVU cohort that included a total of 747 COVID-19 positive, 1223 influenza and 1913 COVID-19 negative patients (Table 1, Figure S1).

### TriNetX External Cohort Dataset

The external validation cohort was obtained from the TriNetX Research network, a cloud-based database resource that provides researchers access to de-identified patient data from networks of healthcare organizations (HCO), mainly from large academic centers and other data providers. The data is directly pulled from electronic medical records and accessible in a de-identified manner. The TriNetX database includes patient-level demographic, vital signs, diagnoses, procedures, medications, laboratory, and genomic data. For this study, we identified COVID-19 and influenza cases using the International Classification of Diseases, 10^th^ revision. We used ICD-10 codes J9.0, J10.0, J09.*, and J10.* for influenza, and U07.1 for COVID-19. Cases from February 1^st^, 2020, to August 13^th^, 2020, were queried and extracted. Due to heterogeneity in reporting of COVID-19 testing (i.e., positive vs. negative; normal vs. abnormal), we refrained from selecting the patients based on their testing results and used diagnostic codes instead. Thus, we were only able to extract COVID-19 positive and influenza positive patients for external validation. Further patients at least 18 years or older at the time of diagnosis and their encounters defined as an encounter occurring within 7days from the date of diagnosis were enforced to be included in the final cohort (n=34,670). Enforcing no missing values corresponding to body temperature resulted in a dataset (n=15,697) that had a total of 6,613 (n=1057 patients) and 9084 (n=2068 patients) encounters diagnosed with COVID-19 and Influenza, respectively (Figure S1). Baseline characteristics of the cohort is described in Supplementary Table 2. Since the TriNetX data is de-identified, Institutional Board Review (IRB) oversight is not necessary.

### Data Preprocessing & Split

Different clinical variables investigated in this study include (a) vital signs i.e., body temperature, heart rate, breathing/respiratory rate, oxygen saturation or SPO_2_ and blood pressure, (b) encounter date/type, (c) reason for visit, and (d) basic demographics information such as Age, Gender, Race, and Ethnicity. Of the features considered, the data elements corresponding to sex and ethnicity were binarized for being “female” and “Hispanic or Latino”, respectively. The race feature was further one-hot encoded to three variables i.e., race_White/Caucasian, race_Black/African American and race_Other. Finally, the date of the encounter was converted to represent a numerical value for month, rather than specific day/year to remove the granularity and capture the seasonal effects of influenza.

### Predictive Model Development using Extreme Gradient Boosting Trees

#### Model Training and Testing

XGBoost, a gradient-boosted tree algorithm, was used to develop classifiers to distinguish between patient encounters with confirmed COVID-19 from that of either influenza patient encounters or other unknown viral infections. Separate models were developed to predict confirmed COVID-19 patient encounters post Feb 1, 2020 from either influenza and/or those that tested negative for COVID-19 (i.e., COVID-19 positive vs influenza, COVID-19 positive vs COVID-19 negative, COVID-19 positive vs influenza vs COVID-19 negative), and then an additional model to test influenza infections from others i.e., patients that were either positive or negative to a COVID-19 test (Influenza vs Others). For the purpose of model training and testing, we only consider data from 01-Feb-2020 until 04-Nov-2020 for COVID-19 positive and negative cohorts and from 01-Jan-2019 to 04-Nov-2020 for the influenza patient cohort. We partitioned the dataset into training and testing sets, using an 80%–20% split (Table S1). The holdout test, or internal validation, set is never seen by the model during training and was used only during performance evaluation.

Gradient-boosted trees were selected due to their ability to model complex nonlinear relationships, while robustly handling outliers and missing values. Gradient-boosted trees fuse the concepts of gradient descent (in the loss function space) and boosting. Simpler tree-based models are built additively like a boosting ensemble, to fit the gradient of the loss function for every data point. Only a fraction of the trees fit to the gradient of the loss per data point in the training set. This is analogous to small steps of gradient descent in the loss function space.

For developing the COVID-19 positive vs negative, Influenza vs Others and COVID-19 vs Influenza XGBoost models, the value of learning rate was set to 0.02 and the total number of trees was 600. The alpha value was set to 0 and lambda was set to 1, i.e., we use L2 regularization. Further, min_child_weight was set to 1, to allow highly specific patterns to be learnt as well. To counter the possibility of overfitting, the max_depth parameter of the XGBoost model was set to 4, as more complex features are learnt with higher depths, leading to poor performance on the unseen data. We used the gain importance metric to decide node splits for the tree estimator, and the objective was set to binary logistic regression. The model outputs the predicted probability, and records whose probability was at least 0.5 were considered as belonging to the positive class, while others were deemed to belong to the negative class. Further, the subsample parameter of the XGBoost algorithm represents the ratio of the records to be sampled before forming a tree and was set to a value of 0.8. Similarly, the colsample_bytree parameter which determines the fraction of columns to be sampled before every tree formation was set to 1, meaning all features are used. Only records with all vitals were considered, however the model does have the ability to make predictions in the absence of certain features. The compute_sample_weight from the sklearn.utils.class_weight library was employed to ensure that each training sample is weighted based on its class, to counter imbalance in the dataset.

For the three-class classifier that discriminates between COVID-19-positive, COVID-19-negative and Influenza encounters, we developed an XGBoost model of 100 trees, with learning_rate set to 0.3 and max_depth fixed at 6. The objective was set to softprob (uses a softmax objective function), to perform multi-class classification. Furthermore, the subsample parameter was set to 1, meaning all samples were used to form trees. No sample weighting was carried out for this model. All other parameters remained the same as the models discussed previously.

#### Model Validation

We also consider an unseen external validation dataset (TriNetX cohort) to evaluate the performance of the model on data that is completely different from what it is accustomed to during training. Patient-wise encounters with no missing vitals were enforced to test and evaluate the performance of the developed models.

#### Model Interpretability using SHAP

To identify the principal features driving the model prediction, SHAP values were calculated. The SHAP method is suitable for the interpretation of complex models such as artificial neural networks and gradient-boosting machines (e.g., XGBoost) ^31,32^. Originating in game theory, SHAP provides model output explanations to answer how does a given prediction change when a particular feature is removed from the model. The resulting SHAP values quantify the magnitude and direction (positive or negative) of a feature’s effect on a given prediction. Thus, SHAP partitions the prediction result of every sample into the contribution of each constituent feature value by estimating differences between model outputs with subsets of the feature space. By averaging across samples, this method helps estimate the contribution of each feature to overall model predictions for the entire dataset. The SHAP is also representative of how important a feature is to the prediction - larger the absolute SHAP value of the feature, the greater its impact on predictions. The direction of SHAP values in force plots represents whether the feature is influential or indicative for the negative or positive class.

#### Performance Evaluation Metrics

The discriminative ability of each of the models developed in predicting patients with influenza from COVID-19 positive and /or negative patients as well as those with a positive COVID-19 test from negative test was evaluated in the hold-out test set and an external validation test by using receiver operator characteristic (ROC) curve analysis. An area under the curve (AUC)□>□0.5 indicated better predictive values. The closer the AUC was to 1, the better the model’s performance was. Additionally, “classification accuracy”, the total number of true positive scores i.e. when predicted values are equal to the actual values given by the attribute positive predictive value (PPV) or “precision” score, “recall” which is the total number of true positive instances among all the positive instances, and “F1 score” the weighted harmonic mean of precision and recall along with negative predictive value (NPV) were estimated to evaluate performance and the overall generalizability of each of the models. Further details on the calculations of different metrics employed are provided in Supplementary methods.

### Statistical analysis

We performed the Shapiro-Wilk test on the entire WVU internal cohort dataset to check for normalcy of the data. We found that all the variables were normally distributed; therefore, we used parametric methods for further statistical analysis. Categorical data are presented as counts (percentages) while continuous data were reported as mean ± standard deviation (SD). To determine the significance of continuous variables between all three different groups (i.e., COVID-19 positive, influenza, and COVID-19 negative) Kruskal-Wallis test with Dunn-Bonferroni correction test was used. The comparison between two of these three groups (COVID-19 positive vs negative, COVID-19 positive vs influenza, influenza vs COVID-19 negative) to determine significance was performed using an independent sample t-test. Chi-square was used for the categorical variable because the expected value for each cell is greater than 5. All statistical analysis was performed using Medcalc for Windows, version 19.5.3 (MedCalc Software, Ostend, Belgium), and python for windows, version 3.8.3. A statistical significance level of p < 0.05 was used for all the tests performed.

## Data Availability

The data that support the findings of this study are available from WVU CTSI & TriNetX but restrictions apply to the availability of these data, which were used under license for the current study, and so are not publicly available. Data is however available from the authors upon reasonable request and with permission of WVU CTSI & TriNetX.

https://github.com/ynaveena/COVID-19-vs-Influenza

## Code availability

The code is made available at https://github.com/ynaveena/COVID-19-vs-Influenza for non-commercial use.

## Acknowledgments

The authors acknowledge Matthew Armistead, Emily Morgan, Maryam Khodaverdi, and Wes Kimble for their support with data collection and providing de-identified datasets.

## Author Contributions

P.P.S and NY envisioned and designed the projects. NY, N.H.K, AR, and SS performed machine learning modeling and analysis. P.P.S., Q.A.H, BP, and PF was involved with clinical interpretation of the findings and writing as well as editing the manuscript. H.B.P and S.S. performed the statistical analysis. All the authors have significantly been involved in the drafting of the manuscript or revising it critically for important intellectual content.

## Competing Interests

The authors declare the following competing interests. Dr. Sengupta is a consultant for HeartSciences, Kencor Health and Ultromics. The remaining authors declare no competing interests.

## Notes

### Competing Interest Statement

The authors declare the following competing interests. Dr. Sengupta is a consultant for Kencor Health and Ultromics. The remaining authors declare no competing interests.

### Funding Statement

This work is supported in part by funds from the National Science Foundation (NSF: # 1920920) National Institute of General Medical Sciences of the National Institutes of Health under (NIH: #5U54GM104942-04). The content is solely the responsibility of the authors and does not necessarily represent the official views of the National Institutes of Health or National Science Foundation.

### Author Declarations

This study protocol was reviewed by the West Virginia University Institutional Review Board and ethical approval was given by WVU IRB.

