## Supplemental Materials for "A Vital Sign-based Prediction Algorithm for Differentiating COVID-19 Versus Seasonal Influenza in Hospitalized Patients"

**Supplemental Information**

**Supplemental Figures**

**Figure S1.** Flowchart showing the study outline and details of the WVU and TrinetX COVID-19 positive, -negative and influenza patient subgroups of considered for this study.

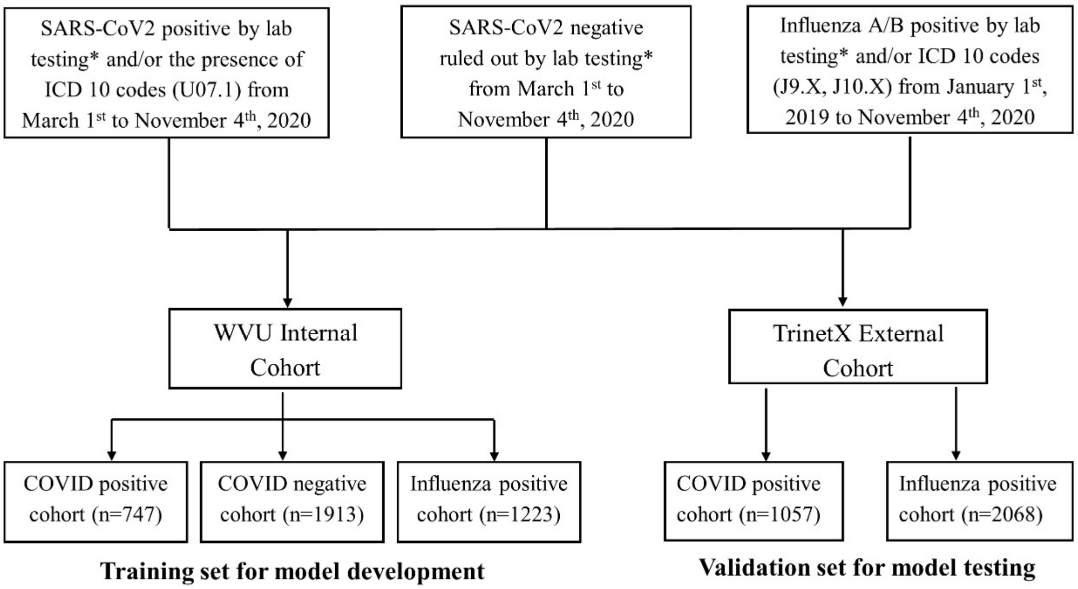


** Confirmed diagnostic testing data is only available for WVU cohort.*

**Figure S2.** SHAP summary plot of the multi-class classifier model that can predict patients that could test positive for Influenza, COVID-19 positive or -negative .

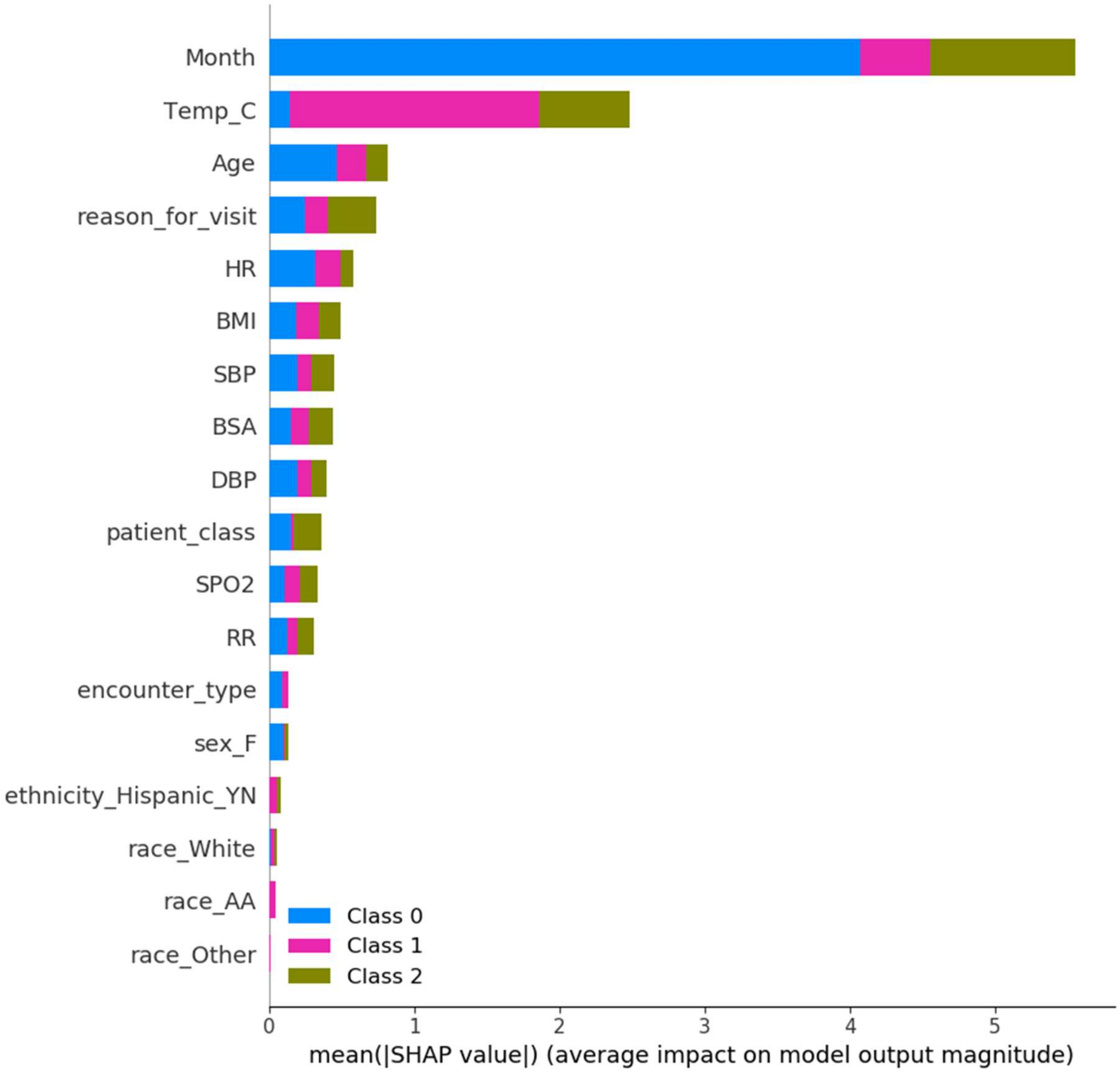

**Figure S3.** The Precision-Recall Curve for the COVID-19-positive vs -negative XGBoost model.

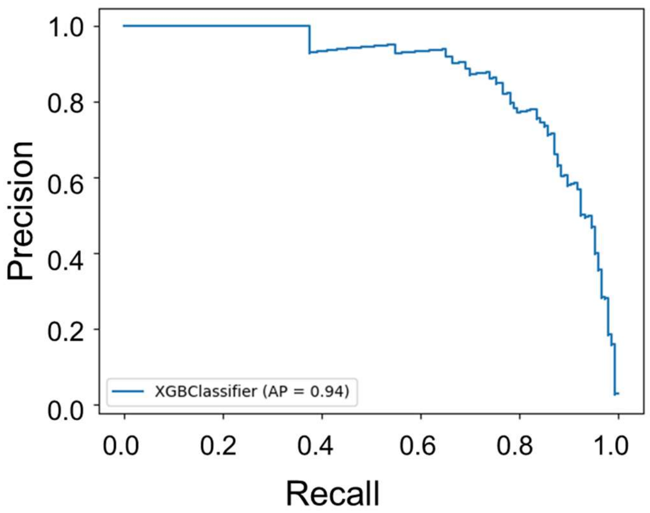

**Figure S4.** Impact of (a) sequential removal of vitals based on feature importance, and (b) including only one vital sign at a time on the predictive model performance. When considering the importance of a feature pertaining to each vital sign in (b), all other vitals were removed from the internal validation set. However, the demographics information was not ablated.

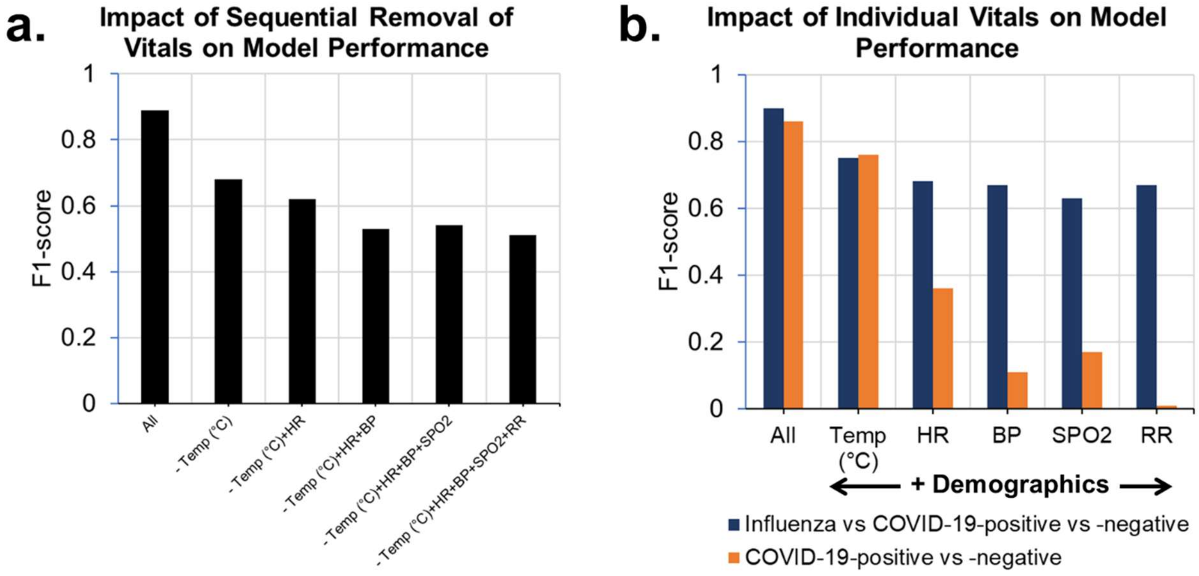

**Supplemental Tables.**

Table S1. Baseline characteristics of the model development/training and model test/internal validation cohort from WVU hospitals.

| Attribute Name | Train | | | Test | | |
|---|---|---|---|---|---|---|
| | COVID-19-pos | Influenza | COVID-19-neg | COVID-19-pos | Influenza | COVID-19-neg |
| Patients Count | 601 | 961 | 1544 | 146 | 262 | 369 |
| Body Mass Index | 31.74 ± 8.96 | 29.75 ± 9.48 | 29.65 ± 9.33 | 30.89 ± 8.33 | 29.58 ± 9.42 | 29.62 ±10.36 |
| Body Surface Area | 2.07 ± 0.35 | 1.93 ± 0.44 | 1.96 ± 0.39 | 2.05 ± 0.35 | 1.91 ± 0.41 | 1.96 ± 0.42 |
| **Sex** | | | | | | |
| Male | 299 | 454 | 740 | 76 | 131 | 176 |
| Female | 302 | 507 | 804 | 70 | 131 | 193 |
| **Ethnicity** | | | | | | |
| Hispanic or Latino | 30 | 29 | 14 | 4 | 14 | 3 |
| Not Hispanic or Latino | 571 | 932 | 1530 | 142 | 248 | 366 |
| **Race** | | | | | | |
| Black or African American | 41 | 77 | 46 | 7 | 18 | 14 |
| White | 513 | 816 | 1437 | 128 | 221 | 340 |
| Others | 47 | 68 | 61 | 11 | 23 | 15 |
| **Age Group** | | | | | | |
| Children and Teens (0 - 20) | 16.16 ± 4.52 (55) | 11.42 ± 4.57 (236) | 12.04 ± 6.09 (105) | 15.88 ± 4.52 (8) | 11.65 ± 4.02 (75) | 9.29 ± 7.22 (18) |
| Adults (21 - 44) | 31.68 ± 7.72 (137) | 31.01 ± 6.59 (279) | 31.83 ± 6.92 (338) | 33.32 ± 7.83 (34) | 31.9 ± 6.96 (75) | 32.79 ± 6.98 (92) |
| Older Adults (45 - 64) | 55.24 ± 5.9 (178) | 54.74 ± 5.86 (222) | 55.38 ± 5.76 (406) | 55.76 ± 6.28 (38) | 54.31 ± 6.02 (60) | 54.23 ± 6.12 (98) |

| | | | | | | |
|---|---|---|---|---|---|---|
| Senior Adults (65+) | 76.52 ± 8.19 (231) | 74.65 ± 7.72 (215) | 77.7 ± 8.24 (673) | 77.35 ± 8.04 (66) | 77.34 ± 7.03 (50)* | 77.2 ± 8.1 (154) |
| **Vitals** | | | | | | |
| Oxygen Saturation (SpO2) | 95.64 ± 4.37 (601) | 96.67 ± 2.8 (961) | 96.28 ± 3.48 (1539) | 95.58 ± 3.49 (146) | 96.81 ± 3.59 (262) | 96.29 ± 5.52 (369) |
| Body Temperature (Celcius) | 36.86 ± 0.6 (601) | 37.18 ± 0.71 (961) | 36.63 ± 0.45 (1539) | 36.85 ± 0.55 (146) | 37.15 ± 0.72 (262) | 36.66 ± 0.5 (369) |
| Diastolic BP | 75.52 ±13.55 (601) | 72.21 ±11.67 (961) | 72.42 ± 13.2 (1539) | 73.51 ±12.97 (146) | 70.41±12.18 (262) | 72.95±12.92 (369) |
| Systolic BP | 127.37±18.8 (601) | 124.26±19.1 (961) | 125.9 ±19.9 (1539) | 126.11 ±19.1 (146) | 122.74±18.2 (262) | 125.93±20.6 (369) |
| Heart Rate | 83.28±15.86 (601) | 93.45±19.47 (961) | 83.04±16.84 (1539) | 80.94 ± 16.2 (146) | 94.73±20.07 (262) | 85.39±18.48 (367)* |
| Respiratory Rate | 18.53 ± 3.6 (601) | 18.85 ± 3.19 (961) | 18.21 ± 3.48 (1539) | 18.59 ± 3.26 (146) | 18.95 ± 3.21 (262) | 18.71 ± 9.42 (368) |

Table S2. Patient demographics of the external validation cohort derived from the TrinetX research network.

| | Overall | COVID-positive | Influenza | p value |
|---|---|---|---|---|
| **N** | 3125 | 1057 | 2068 | |
| **Encounters** | 15,697 | 6613 | 9087 | |
| **Sex** | | | | 0.0001* |
| Male | 1528 | 568 (53.7) | 960 (46.4) | |
| Female | 1597 | 489 (46.3) | 1108 (53.6) | |
| **Ethnicity** | | | | 0.0001* |
| Hispanic or Latino | 74 | 41 (3.9) | 33 (1.6) | |
| Not Hispanic or Latino | 3051 | 1016 (96.1) | 2035 (98.4) | |
| **Race** | | | | <0.0001* |
| African American | 1491 | 523 (49.5) | 968 (46.8) | |
| White | 1254 | 307 (29.0) | 947 (45.8) | |
| Others | 380 | 227 (21.5) | 153 (7.4) | |
| **Overall Age Group** | 57.15±17.12 | 57.22±16.64 | 57.11±17.36 | 0.87 |
| Children and Teens (0 - 20) | 19.27±0.70 | 18.75±0.50 | 19.39±0.70 | 0.10 |
| | 22 | 4 (0.4) | 18 (0.9) | |
| Adults (21 - 44) | 33.33±6.73 | 33.30±6.58 | 33.35±6.81 | 0.93 |
| | 719 | 234 (22.1) | 485 (23.5) | |
| Older Adults (45 - 64) | 55.88±5.51 | 55.83±5.57 | 55.91±5.48 | 0.80 |
| | 1214 | 435 (41.2) | 779 (37.7) | |
| Senior Adults (65+) | 74.47±6.94 | 74.25±6.78 | 74.58±7.02 | 0.45 |
| | 1125 | 373 (35.3) | 752 (36.4) | |
| **Vitals** | | | | |
| Oxygen Saturation (SpO2) | 95.94±2.49 | 95.80±2.71 | 96.08±2.23 | 0.03* |
| | 1580 | 796 (50.4) | 784 (49.6) | |
| Body Temperature (°Farenheight) | 98.57±1.28 | 98.55±0.98 | 98.58±1.46 | 0.2 |
| | 15697 | 6613 (42.1) | 9084 (57.9) | |
| Diastolic BP | 73.09±11.13 | 73.87±10.72 | 72.24±11.50 | <0.0001* |
| | 9435 | 4910 (52.0) | 4525 (48.0) | |
| Systolic BP | 126.44±18.50 | 127.10±17.86 | 125.73±19.15 | 0.0003* |
| | 9436 | 4911 (52.0) | 4525 (48.0) | |
| Heart Rate | 88.49±15.98 | 86.94±15.45 | 89.53±16.25 | <0.0001* |
| | 13833 | 5583 (40.4) | 8250 (59.6) | |
| Respiratory Rate | 20.12±4.38 | 20.89±5.01 | 19.56±3.76 | <0.0001* |
| | 15691 | 6611 (42.1) | 9080 (57.9) | |

Values are counts (%) or mean ± standard deviation.
*p < 0.05 between COVID-Positive and Influenza group. P values were calculated using independent t test where mean is reported and Chi squared where frequencies are reported.

**Table S3.** Assessment of performance of multi-class classification model at identifying Influenza or COVID-19 amongst patients presenting to or admitted at the hospital in a held-out 20% test set.

| Performance Measure | Influenza vs Rest | COVID-19-positive vs Rest | COVID-19-negative vs Rest |
|---|---|---|---|
| AUC | 0.983 | 0.974 | 0.971 |
| Accuracy | 0.941 | 0.945 | 0.916 |
| F1 Score | 0.912 | 0.837 | 0.915 |
| Sensitivity (Recall) | 0.912 | 0.753 | 0.882 |
| Specificity | 0.955 | 0.989 | 0.953 |
| PPV (Precision) | 0.912 | 0.940 | 0.951 |
| NPV | 0.955 | 0.946 | 0.885 |
| FPR | 0.045 | 0.011 | 0.048 |
| FDR | 0.087 | 0.060 | 0.049 |
| FNR | 0.087 | 0.247 | 0.118 |

**Table S4.** Chief complaint or reason for visit among COVID-positive, -negative and influenza patients.

| | COVID Positive | Influenza | COVID Negative |
|---|---|---|---|
| **Total (%)** | **747** | **1223** | **1913** |
| Respiratory (SOB, Cough, URI/Flu/COVID like symptoms) | 339 (51.21) | 625 (55.83) | 515 (34.85) |
| Fever | 82 (12.39) | 218 (18.78) | 81 (5.42) |
| GI (nausea, vomiting, diarrhea, abdominal pain, GI bleed) | 60 (9.06) | 71 (6.12) | 143 (9.57) |
| Chest pain or other heart problem | 24 (3.63) | 50 (4.31) | 103 (6.89) |
| CNS (alter mental status, dizziness, syncope, headache, stroke, fall, confusion) | 55 (8.31) | 45 (3.88) | 222 (14.85) |
| Systemic (generalized fatigue, lethargy, weakness, bodyache, hypo/hyperglycemia lightheadedness, hypotension) | 51 (7.70) | 59 (5.08) | 89 (5.95) |
| Other (skin problem, urinary problem, pain, injury/trauma, OBGyn problems, ENT problems, medical evaluation, or abnormal tests evaluation, etc.) | 51 (7.70) | 50 (4.31) | 342 (22.88) |
| Unknown | 85 (11.38) | 62 (5.07) | 418 (21.85) |

SOB - Shortness of breath; GI - Gastrointestinal; CNS - Central Nervous System; OBGyn - Obstetrics and Gynecology; ENT - ear, nose, and throat

**Table S5.** Assessment of the performance of Influenza vs COVID-19-positive and Influenza vs Other predictive models at identifying Influenza or COVID-19 amongst patients presenting to or admitted at the hospital in the TriNetX external validation test set. The datasets evaluated below enforces three conditions: (a) no missing values are allowed for body temperature (n=15,697) and (b) no missing values are allowed for both heart rate and body temperature (n=13,834) and (c) no missing values in any of the vital signs (n=1,340).

| **Performance Measure** | **Influenza vs COVID-19-positive** | | | **Influenza vs Others** | | |
|---|---|---|---|---|---|---|
| | **Body Temperature** | **Body Temperature + Heart rate** | **Enforcing no missing Vitals** | **Body Temperature** | **Body Temperature + Heart rate** | **Enforcing no missing vitals** |
| AUC | 0.923 | 0.925 | 0.943 | 0.923 | 0.924 | 0.962 |
| Accuracy | 0.852 | 0.858 | 0.88 | 0.801 | 0.798 | 0.86 |
| F1 Score | 0.827 | 0.823 | 0.89 | 0.807 | 0.798 | 0.86 |
| Sensitivity (Recall) | 0.842 | 0.845 | 0.86 | 0.682 | 0.669 | 0.77 |
| Specificity | 0.860 | 0.867 | 0.90 | 0.989 | 0.989 | 0.98 |
| PPV (Precision) | 0.814 | 0.811 | 0.92 | 0.990 | 0.989 | 0.99 |
| NPV | 0.882 | 0.892 | 0.83 | 0.664 | 0.669 | 0.76 |
| FPR | 0.141 | 0.133 | 0.10 | 0.011 | 0.011 | 0.02 |
| FDR | 0.187 | 0.189 | 0.08 | 0.010 | 0.011 | 0.02 |
| FNR | 0.158 | 0.155 | 0.14 | 0.318 | 0.331 | 0.23 |

### Supplemental Methods.

#### Evaluation Metrics

The following metrics were used to evaluate the performance of various models developed as part of this study. One of the most determined statistical measures is Sensitivity (also known as recall, hit rate or true positive rate TPR). Sensitivity measures the proportion of actual positives that are correctly identified as positives: TP / (TP + FN). Specificity, also known as selectivity or true negative rate (TNR), measures the proportion of actual negatives that are correctly identified as negatives: TN / (FP + TN). The Positive Predictive Value (PPV = TP / (TP + FP)), also known as Precision and the Negative Predictive Value (NPV = TN / (TN + FN)) are measures of the performance of a diagnostic test. The False Positive Rate (FPR = FP / (FP + TN)) or fall-out is the ratio between the number of negative events incorrectly categorized as positive (false positives) and the total number of actual negative events (regardless of classification). The False Negative Rate (FNR= FN / (FN + TP)) measures the proportion of the individuals where a condition is present for which the test result is negative. Accuracy (ACC) is a measure of all correctly predicted observations and defined as (TP + TN) / (TP + TN + FP + FN). The F1 Score is a measure of a test's accuracy, defined as the harmonic mean of precision and recall i.e., 2TP / (2TP + FP + FN).
